# High Brain-Derived Neurotrophic Factor (BDNF) and Low Psychological Flexibility and associate with Fatigue symptoms

**DOI:** 10.1101/2022.03.15.22271536

**Authors:** Nalinee Yingchankul, Siriporn Chattipakorn, Patama Gomutbutra

## Abstract

**Background:** Recent studies showed that enhancing psychological flexibility could improve fatigue interference. Brain-Derived Neurotrophic Factor (BDNF), Heart Rate Variability (HRV), and Cortisol were proposed to involve biomarkers in psychological flexibility. Our study aims to explore the association of fatigue with psychological flexibility and related biomarkers.

**Method:** A cross-sectional study gathered data from a baseline characteristic mindful volunteer. Each participant was self-evaluated with the questionnaire of fatigue and psychological flexibility. The participants were evaluated potential biomarkers related to psychological flexibility including HRV, serum cortisol, and BDNF within one week after responding to the questionnaire.

**Results:** The 47 healthy females including 22 nurses and 25 occupational therapy students, mean age 29.70 ± 12.55 years. The prevalence of fatigue is 38.30%. The multivariate analysis showed the independent factors associated with fatigue including negative psychological flexibility (OR 1.31, p=0.03) and high BDNF (OR 1.33, p=0.05).

**Conclusion:** Our study found that psychological flexibility and high BDNF was independent factors associate with fatigue. This result provide insight that intervention that increase either psychological flexibility may prevent fatigue symptoms. The high BDNF may reflex the adaptive response of fatigue person and may be potential biomarkers for detecting early fatigue conditions.

## Introduction

Fatigue is one of the most common non-specific symptoms in healthy young adult females with psycho-pathophysiology still being under-investigated. Fatigue is a subjective perception of lacking energy or tiredness leading to a decreased capacity for physical and mental capacity to cope with stress(Berrios, 1990). Despite without lethal pathology, fatigue significantly impacts the quality of life and work productivity. A study in Sweden showed females aged 20-35 had higher mental fatigue comparing to older females and males of all ages(Engberg et al., 2017). The high prevalence of fatigue in this young adult female was widely explained by stress from work-life(Skinner & Dorrian, 2015). However, a study in university students also found a higher occurrence of chronic fatigue in female students(Bouloukaki et al., 2017). Therefore, the explanation may be beyond the nature of work but rather a psychophysiology pathway.

Improving psychological flexibility to prevent or reduce mental stress-related fatigue was one of the interesting therapeutic concepts. Psychological flexibility was purposed as an antidote to stress-induced psychosomatic symptoms. The psychophysiology definition of stress is *“a notable and persistent condition in which an organism is exposed to risk factors, which tend to alter its balance or homeostasis”*. Meanwhile, psychological flexibility is defined as “*the acceptance of our own thoughts, emotions and acting on long-term values rather than short-term impulses, thoughts, and feelings that are often linked to experiential avoidance and a way to control unwanted inner events”*. Therefore, it is intuitive that psychological flexibility and adaptive repertoire are crucial in a healthy response to stress(Yu et al., 2020). There were studies found in association with interventions that aimed to improve psychological flexibility, like acceptance and commitment therapy or mindfulness-based therapy, to improve fatigue interference(Hayes et al., 2006; Ramaci et al., 2019). The also improving depression and sleep quality suggested these components likely were either mediators or effect modifiers in the effect of the intervention on fatigue(Kato, 2016).

To date, there is no gold standard to measure psychological flexibility, although there are some well-validated questionnaires (e.g., Acceptance and Action Questionnaire)(Hayes et al., 2006). There were proposed objective physiologic biomarkers associated with psychological flexibility including Heart Rate Variability (HRV)(Walker et al., 2017). Besides, there is an increasing interest role of Brain-Derived Neurotrophic Factor (BDNF), as a cognitive flexibility biomarker. HRV is an index of a healthy autonomic nervous system function called sympathovagal balance. An increase in HRV reflects the variability in time elapsed between heartbeats which indicated healthy stress adaptability. A study in neurofibromatosis patients showed lower HRV associated with psychological flexibility and difficult pain management (Allen et al., 2018). BDNF, a neurotrophic protein well recognized in the preclinical study as having important roles in synaptic plasticity in brain areas that mediate executive function as, the prefrontal cortex (PFC) and learning new knowledge as the hippocampus. In rodents, PFC BDNF rapidly increases and peaks in early adolescence and then gradually decreases to adult levels(Xu et al., 2016). A previous study found an association between lower BDNF and maladaptive psychological diseases like post-traumatic stress disorder, depression, schizophrenia in adolescents (Autry & Monteggia, 2012). However, a study showing an association between BDNF, psychological flexibility, and fatigue has not been done.

Our study aims to evaluate 1) The association of fatigue and psychological flexibility and the potential mediators as depression and sleep quality among apparent healthy females. 2) The association of fatigue and psychological flexibility physiologic markers as HRV, Cortisol, and BDNF. Our study conceptual framework is depicted in ***Figure1***.

**Figure 1:**
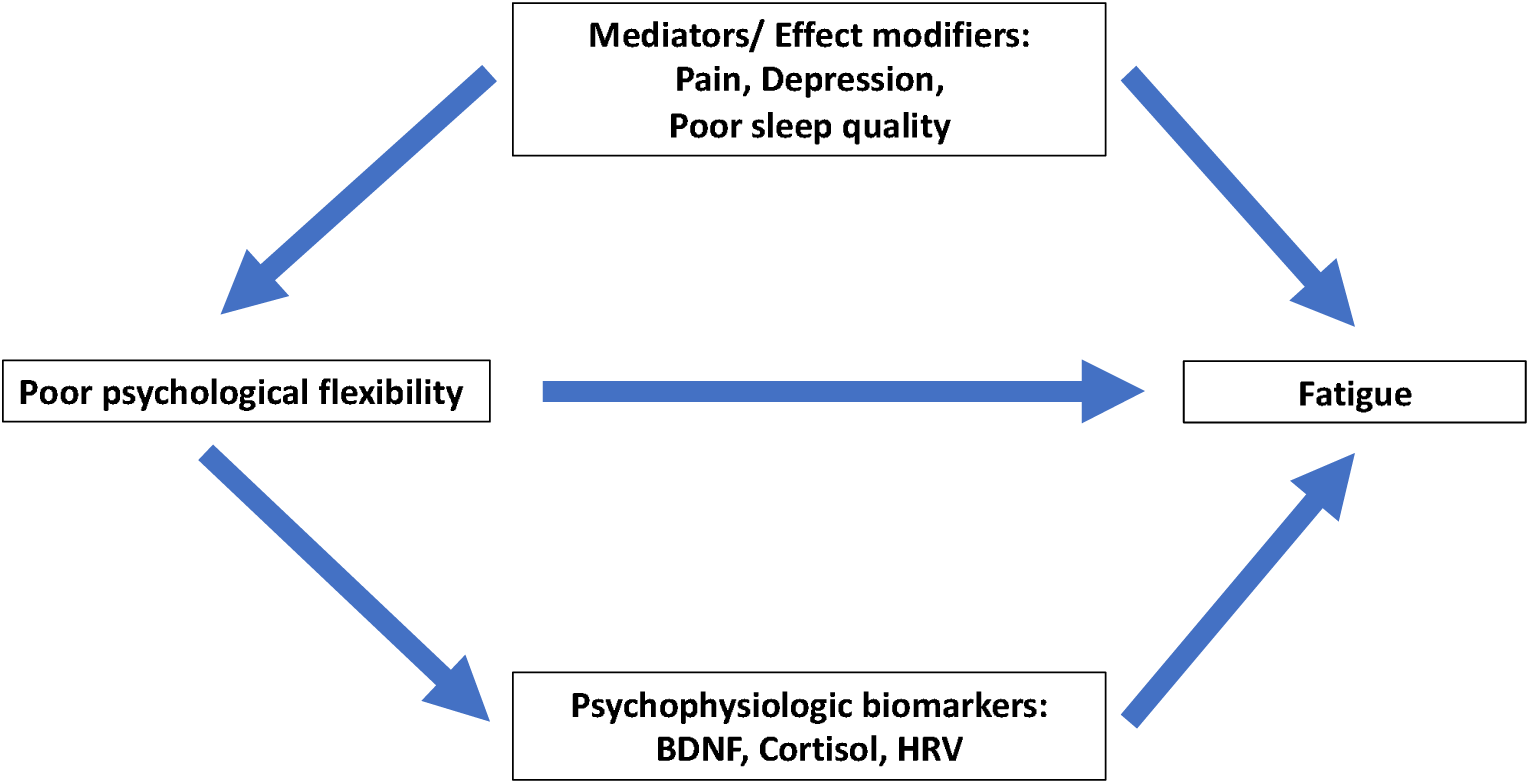
The hypothesized association (Abbreviation: HRV = Heart Rate Variability, BDNF = Brain Derived Neurotrophic Factor)

## Methods

A cross-sectional pilot study was done between November 2018 – March 2020 in Maharaj Nakorn Chiang Mai Hospital. The inclusion criteria were adults ages over 18 years old, can read and self-answer questionnaire in both English and Thai language, allowed permission to collect data from their blood results. Calculating the sample size for estimating an infinite population was referenced from a previous study that showed the prevalence of fatigue is 25%(Cullen et al., 2002). The sample size will be estimated on this value by one - sample comparison of a proportion assigned a two-sided alpha level of 0.05 and a power of 80%. As a result of sample size calculation, the sample size of this research should be at least 19 samples. Data collection was done by self-reporting questionnaires and laboratory examination. There are 6 parts which are 1. Demographic data, 2. Psychological flexibility questions; 13 items of negative psychological flexibility questions. The score for each item is sorted according to severity; 0 = never or without, 3 = most have or most severe then calculate the total score (total 39) as shown detail in appendix. 3. Fatigue questionnaire; 3 items of fatigue questionnaire. 0 = never or without, 3 = most have or most severe then calculate the total score (total 9). 4. Depression screening questionnaire (PHQ-9), 5. Sleep quality questionnaire (PSQI), 6. Digit span, 7. Pain; 0 = never or without, 3 = most have and 8. Laboratory measurement; Heart Rate Variability (HRV), serum BDNF, serum morning glucose, serum morning cortisol. This study was approved by the Maharaj Nakorn Chiang Mai Hospital Research ethics committee. Research number 353/2561. The analytical statistical calculation was performed using Stata version 12 with frequency, mean, chi-squared test, Fisher’s exact test, t-test, and multivariate logistic regression. The negative psychological flexibility questions had content validation by experts, there are good levels of internal reliability, with Cronbach’s alpha coefficients being 0.8309.

## Results

From the demographic data of 47 female volunteers in this study are 18 nurses, 4 nurse assists, and 25 occupational therapy students. The average age is 29.70 ± 12.55 years. All participants were healthy or did not have an uncontrolled disease; 12.77% have chronic illnesses that require continuous medication intake. 74.47% are single. 25.53% have night shifts. From the subjective measurement outcome in this study, the prevalence of fatigue symptoms was 38.30%. The average negative psychological flexibility score was 11.55 ± 5.73. The average PSQI score is 6.89 ± 2.99. 59.57% were scored poor sleep quality from PSQI but only 23.40% of the self-evaluated had poor sleep quality. 19.57% had depression from the PHQ-9 questionnaire; the mean score was 4.78 ± 3.41. Mean morning serum cortisol level was 10.19 ± 5.59 μg/dl. Mean serum BDNF was 6.39 ± 3.64 ng/ml, HRV results; mean LF/HF ratio was 1.54 ± 0.49, and mean RMSSD was 30.40 ± 11.69, as shown in ***Table1***.

**Table1:** Demographic data and measurement outcome.

| **Factors** |  | **(N=47)** |
| --- | --- | --- |
| Female sex | n (%) | 47 (100) |
| Age (year) | Mean ± SD | 29.70 ± 12.55 |
| Occupation |  |  |
| Register nurse | n (%) | 18 (38.30) |
| Nursing assistant | n (%) | 4 (8.51) |
| University student | n (%) | 25 (53.19) |
| Education |  |  |
| Master | n (%) | 4 (8.51) |
| Bachelor | n (%) | 14 (29.79) |
| University student | n (%) | 25 (53.19) |
| High school | n (%) | 4 (8.51) |
| Marital status |  |  |
| Single | n (%) | 35 (74.47) |
| Married | n (%) | 12 (25.53) |
| Have controlled underlying disease | n (%) | 6 (12.77) |
| Night shift | n (%) | 12 (25.53) |
| Digit span score (missing 7) | Mean ± SD | 14.22 ± 5.81 |
| Forward | Mean ± SD | 9.50 ± 3.50 |
| Backward | Mean ± SD | 5.08 ± 2.10 |
| Sleep quality (PSQI score) | Mean ± SD | 6.89 ± 2.99 |
| Poor sleep quality from PSQI score | n (%) | 28 (59.57) |
| Self-evaluate poor sleep quality | n (%) | 11 (23.40) |
| Depression score (PHQ-9) (missing 1) | Mean ± SD | 4.78 ± 3.41 |
| Positive depression (PHQ-9 ≥ 7) (missing 1) | n (%) | 9 (19.57) |
| Pain score (total score 3) | Mean ± SD | 0.98 ± 0.87 |
| Have pain (score ≥ 1 of 3) | n (%) | 32 (68.09) |
| Fatigue score | Mean ± SD | 2.21 ± 1.52 |
| Have fatigue (total score ≥ 3 of 9) | n (%) | 18 (38.30) |
| Negative psychological flexibility score (total score 39) | Mean ± SD | 11.55 ± 5.73 |
| Morning serum glucose (mg/dl) | Mean ± SD | 89.74 ± 15.64 |
| Morning serum cortisol (μg/dl) | Mean ± SD | 10.19 ± 5.59 |
| Serum BDNF (ng/ml) | Mean ± SD | 6.42 ± 3.72 |
| Heart Rate Variability (HRV) |  |  |
| • Frequent domain |  |  |
| Low frequency (LF) | Mean ± SD | 22.94 ± 10.06 |
| High frequency (HF) | Mean ± SD | 16.30 ± 7.94 |
| LF/HF ratio | Mean ± SD | 1.54 ± 0.49 |
| • Time domain |  |  |
| SDNN | Mean ± SD | 54.83 ± 15.72 |
| RMSSD | Mean ± SD | 30.40 ± 11.69 |

In the bivariate analysis of factors related to the fatigue symptom, five factors were significantly associated with fatigue; negative psychological flexibility, sleep quality, depression, pain symptom, and BDNF. The fatigue group had more negative psychological flexibility scores than the non-fatigue group significantly with a mean score of 15.44 ± 5.81 and 9.14 ± 4.19 respectively(p<0.05). The fatigue group that had poor sleep quality defined by PSQI had more fatigue than those without fatigue, 77.78% and 48.28% respectively, with statistical significance (p<0.05). Groups with depression had more fatigue than those without fatigue, 41.18% to 6.90% respectively, with statistical significance (p<0.05). Groups with pain symptoms had more fatigue than those without fatigue, a mean score of 1.39 ± 1.04 and 0.72 ± 0.65 respectively, with statistical significance (p<0.05). The fatigue group had more BDNF levels than the non-fatigue group significantly (p<0.05); mean score 7.60 ± 3.46 and 5.69 ± 3.74 respectively. as shown in ***Table2***.

**Table2:** Bivariate analysis factors associated with fatigue.

| Factors |  | Fatigue<br>(N=18) | No fatigue<br>(N=29) | p-value |
| --- | --- | --- | --- | --- |
| Age (year) | Mean ± SD | 31.61 ± 13.40 | 28.52 ± 2.24 | 0.42 |
| Occupation | n (%) |  |  | 0.74 |
| Register nurse |  | 8 (44.44) | 10 (34.48) |  |
| Nursing assistant |  | 1 (5.56) | 3 (10.34) |  |
| University student |  | 9 (50.00) | 16 (55.17) |  |
| Education | n (%) |  |  | 0.72 |
| Master |  | 1 (5.56) | 3 (10.34) |  |
| Bachelor |  | 7 (38.89) | 7 (24.14) |  |
| University student |  | 9 (50.00) | 16 (55.17) |  |
| High school |  | 1 (5.56) | 3 (10.34) |  |
| Married | n (%) | 6 (33.33) | 6 (20.69) | 0.33 |
| Have underlying disease | n (%) | 3 (16.67) | 3 (10.34) | 0.66 |
| Night shift (missing 2) | n (%) | 4 (22.22) | 8 (27.59) | 0.74 |
| Digit span (missing 7) | Mean ± SD | 14.38 ± 5.79 | 14.12 ± 5.93 | 0.89 |
| Forward | Mean ± SD | 9.50 ± 3.71 | 9.50 ± 3.44 | 1.00 |
| Backward | Mean ± SD | 4.88 ± 2.28 | 5.21 ± 2.02 | 0.63 |
| Sleep quality (PSQI score) | Mean ± SD | 8.39 ± 2.95 | 5.97 ± 2.67 | <0.01* |
| Poor sleep quality | n (%) | 14 (77.78) | 14 (48.28) | <0.05* |
| Self-evaluate poor sleep quality | n (%) | 6 (33.33) | 5 (17.24) | 0.21 |
| Depression (PHQ-9 score) (missing 1) | Mean ± SD | 6.53 ± 4.33 | 3.76 ± 2.23 | <0.01* |
| Depression (PHQ-9 ≥ 7) (missing 1) | n (%) | 7 (41.18) | 2 (6.90) | <0.01* |
| Pain score | Mean ± SD | 1.39 ± 1.04 | 0.72 ± 0.65 | <0.01* |
| Negative psychological flexibility score | Mean ± SD | 15.44 ± 5.81 | 9.14 ± 4.19 | < 0.01* |
| Morning serum glucose (mg/dl) | Mean ± SD | 88.06 ± 6.78 | 90.79 ± 19.27 | 0.57 |
| Morning serum cortisol ($\mu\text{g/dl}$) | Mean ± SD | 10.31 ± 4.66 | 10.12 ± 6.19 | 0.91 |
| Serum BDNF (ng/ml) | Mean ± SD | 7.60 ± 3.46 | 5.69 ± 3.74 | 0.04** |
| Heart Rate Variability (HRV) |  |  |  |  |
| • Frequent domain |  |  |  |  |
| Low frequency (LF) | Mean ± SD | 22.24 ± 9.71 | 23.38 ± 10.42 | 0.71 |
| High frequency (HF) | Mean ± SD | 15.65 ± 5.22 | 16.71 ± 9.30 | 0.66 |
| LF/HF ratio | Mean ± SD | 1.49 ± 0.56 | 1.57 ± 0.47 | 0.63 |
| • Time domain |  |  |  |  |
| SDNN | Mean ± SD | 54.39 ± 11.23 | 55.10 ± 18.14 | 0.88 |
| RMSSD | Mean ± SD | 29.61 ± 7.29 | 30.89 ± 13.84 | 0.72 |

\*p-value $\leq 0.05$ : statistically significant with null hypothesis the difference of mean equal 0
\*\* p-value $\leq 0.05$ : statistically significant with null hypothesis the mean of fatigue group is more than non-fatigue mean fatigue score of total $9 \pm \text{SD} = 3.83 \pm 0.92$ in fatigue groups, $1.21 \pm 0.73$ in non-fatigue groups

The factors that show significant association with fatigue from bivariate analysis including negative psychological, PHQ9, PSQI, pain score, and BDNF are included in the multivariable analysis model. The results showed that negative psychological flexibility and BDNF are the two independent factors associate with fatigue. Those that had negative psychological flexibility displayed fatigue 1.31 times over those who did not with statistical significance (p=0.03). The participants who had high BDNF levels displayed more fatigue symptoms over than who had not 1.33 times over with statistical significance (p=0.05). The association of PHQ9, PSQI, and pain score becomes not significant as shown in ***Table3***.

**Table3:** The multivariable logistic regression analysis for factors associated with fatigue.

| Factors | Odd ratio | 95%CI | p-value |
| --- | --- | --- | --- |
| Negative psychological flexibility | 1.31 | 1.03 -1.68 | 0.03* |
| BDNF | 1.33 | 1.01-1.78 | 0.05* |
| Pain | 3.43 | 0.89-13.11 | 0.07 |
| PHQ9 | 1.37 | 0.95-1.98 | 0.09 |
| PSQI | 1.25 | 0.94-1.69 | 0.12 |

\* p-value $\leq 0.05$ : statistically significant with null hypothesis the odd ration is not equal 1

The correlational test is done to explore the underline confounder or mediator potential. ***Figure 2*** shows the graphs and Pearson and spearman correlation of negative psychological flexibility, serum BDNF level with PHQ9, PSQI, and pain score. There is no significant correlation between negative psychological flexibility with serum BDNF (r 0.023, p=0.68). There is no significant correlation of BDNF to PHQ9 (r -0.018, p=0.93), PSQI (r 0.202, p=0.23) and pain score (r 0.04, p=0.78). However, there is a strong correlation between negative psychological flexibility and PHQ9 (r 0.514, p<0.01). There is no statistically significant correlation between negative psychological flexibility with PSQI (r 0.222, p= 0.14) and pain (r 0.100, p=0.53).

**Figure 2:**
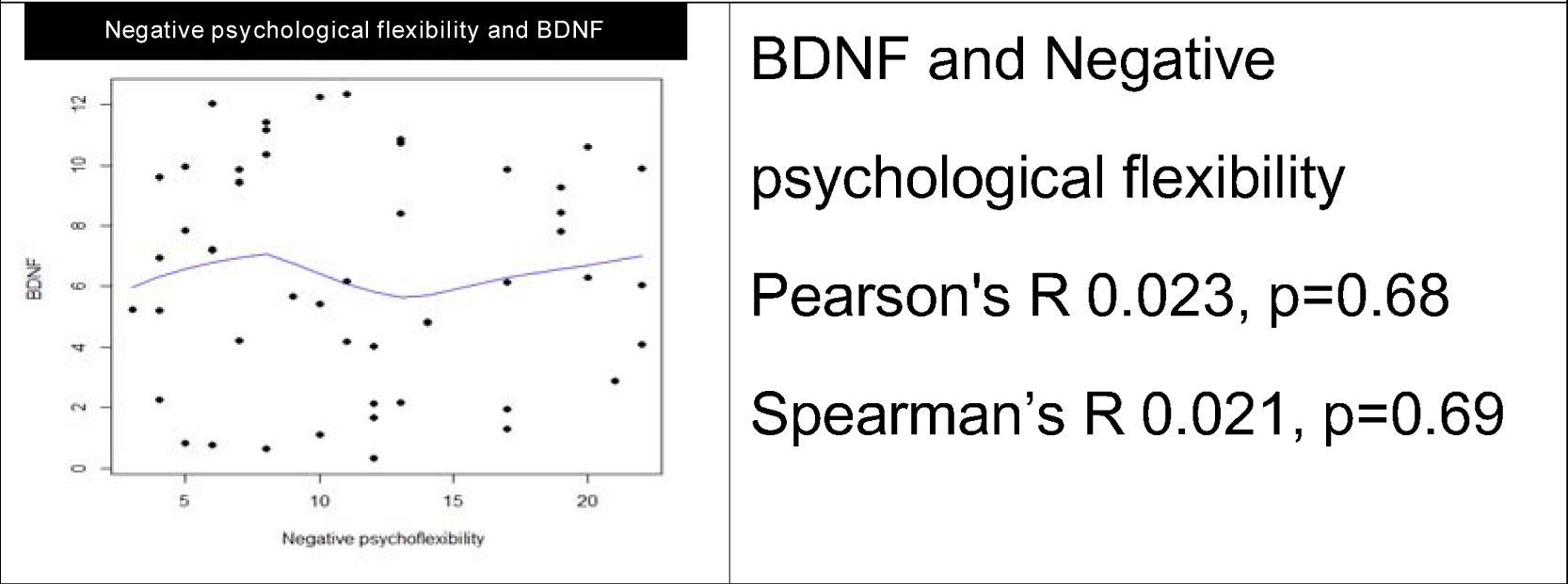
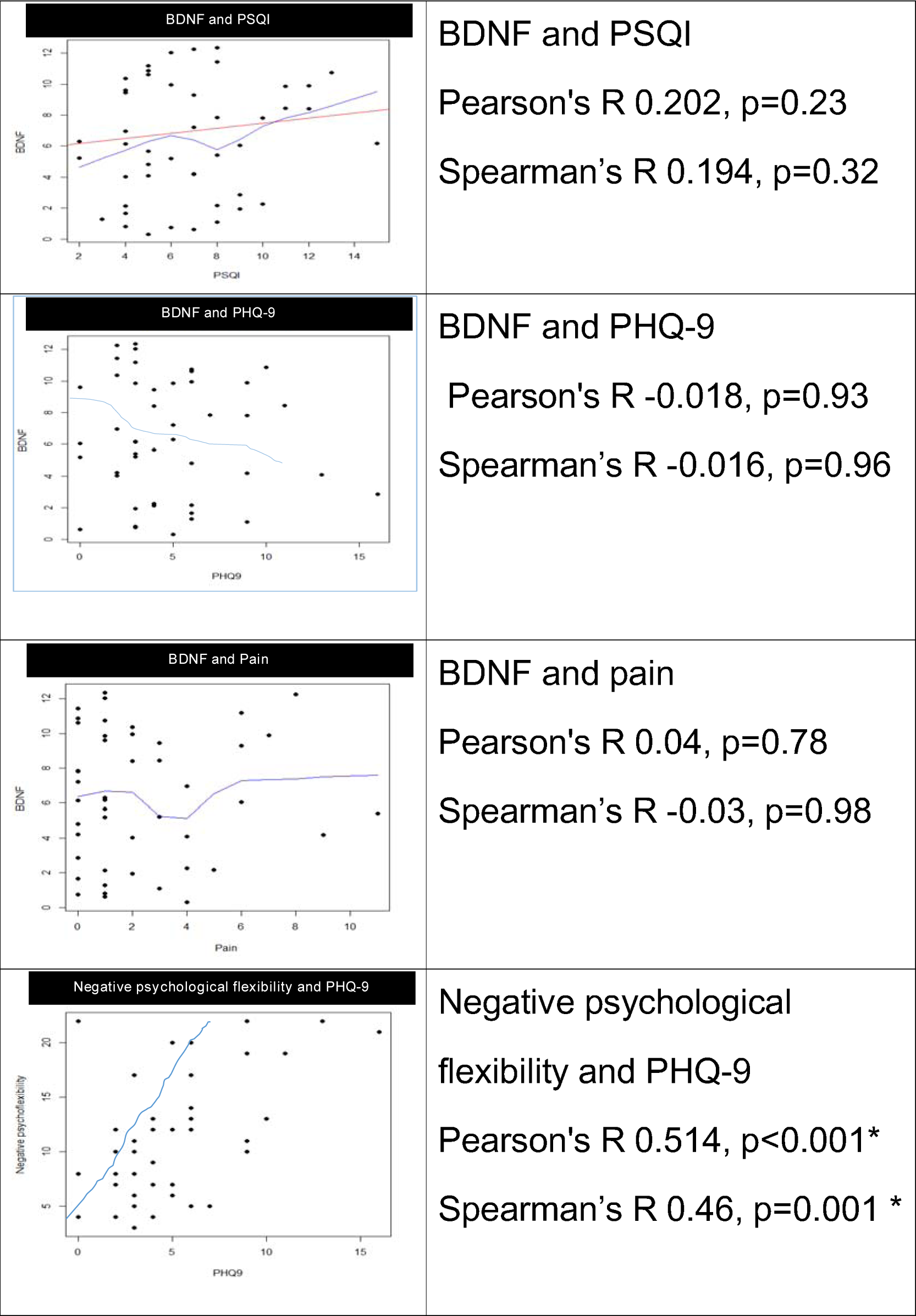
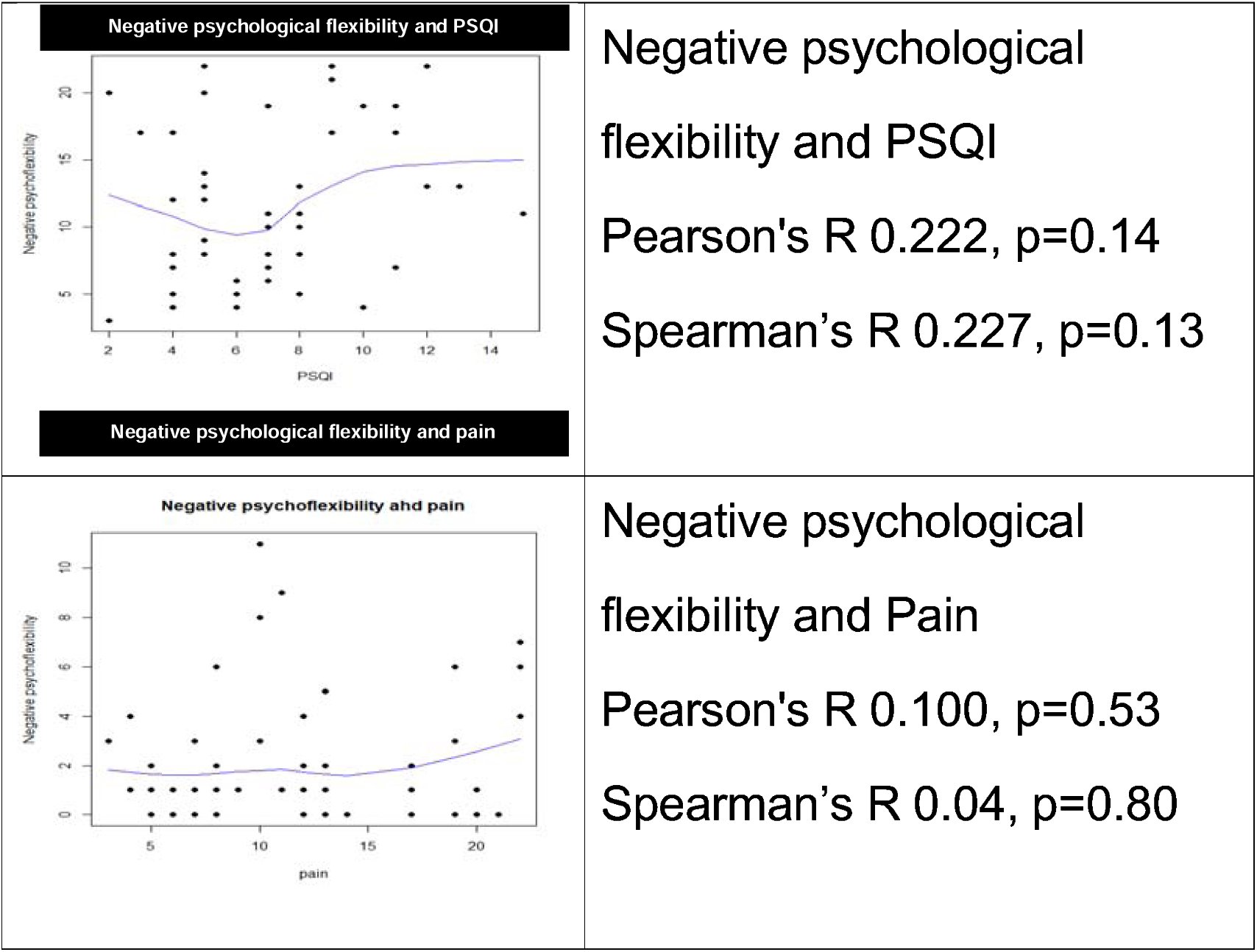
The graph showing the correlation of Negative psychological flexibility, BDNF with PSQI, PHQ9 and pain score.

## Discussion

From the objective, this study showed the significant association of fatigue and psychological flexibility with serum BDNF level among healthy female. The prevalence of fatigue in this study is 38.30%. A previous study found the prevalence of fatigue was 17.2% for the general adult population in Japan(Aritake et al., 2015), 22% in the working population of Dutch companies. A study was done in a primary care clinic in an urban area which showed 27% of the population was concerned about unusual daily living with fatigue for at least 6 months(Bültmann et al., 2002). Our study had higher fatigue prevalence which may be due to participants’ characteristics of a high-stress job and being of female sex along with living in urban areas, corresponding to a systematic review in Bahrain, Egypt, Jordan, Lebanon, Palestine, Saudi Arabia, and Yemen that showed 20 - 81% of health care professionals had emotional exhaustion using Maslach Burnout Inventory assessment(Elbarazi et al., 2017). In which the prevalence is as high as this study. It is possible that subjective fatigue, because it was without signs or complete criteria, was not diagnosed in patients. The difference in criteria for diagnosing fatigue may be the cause of the difference in prevalence. This result suggests that there are quite a lot of people who have symptoms of fatigue and suffer from it. This should be studied further with high sensitivity criteria to diagnose and other relevant factors to help heal these groups.

Our study identifies psychological flexibility as significantly associated with fatigue. A high score of negative psychological flexibility score increases the risk of fatigue 1.31 times. Corresponding to the previous study showed higher positive psychological flexibility had an inverse relationship with fatigue and occupational burnout(Kuba et al., 2019). Therefore, there is need for further studies on the intervention that will help increase the positive psychological flexibility or decrease negative psychological flexibility to reduce fatigue conditions.

BDNF level is an independent factor associate with fatigue, unlike previous evidence suggesting that BDNF is low in severe and chronic fatigue (Saligan et al., 2015). It may be due to the difference of character of the population. In this study, it was a healthy population that had early fatigue, or unqualified fatigue. But in the previous study, it was a population with late or severe fatigue. It is also notable that BDNF has not shown a significant correlation with negative psychological flexibility, depression, poor sleeping, or pain. Therefore, it is interesting if BDNF may link psychophysiology adaptation to stress.

Depression measured by high PHQ9 and pain score are possible confounding variables while poor sleep indicated by high PSQI score is a mediator of the association of negative psychological flexibility and fatigue. These factors made a significant difference between the fatigue and non-fatigued group in the bivariate analysis but the associations disappear after adjusting with negative psychological flexibility and BDNF. The high correlation of negative psychological flexibility and depression indicates the potential confounding. In other words, a psychological flexibility questionnaire could detect depressive symptoms that would underline the cause of fatigue. Meanwhile, poor sleep and pain are likely to be mediators. People with low psychological flexibility character are at risk to develop poor sleep and pain symptoms. The association model from our study is shown in ***Figure 3***

**Figure 3:**
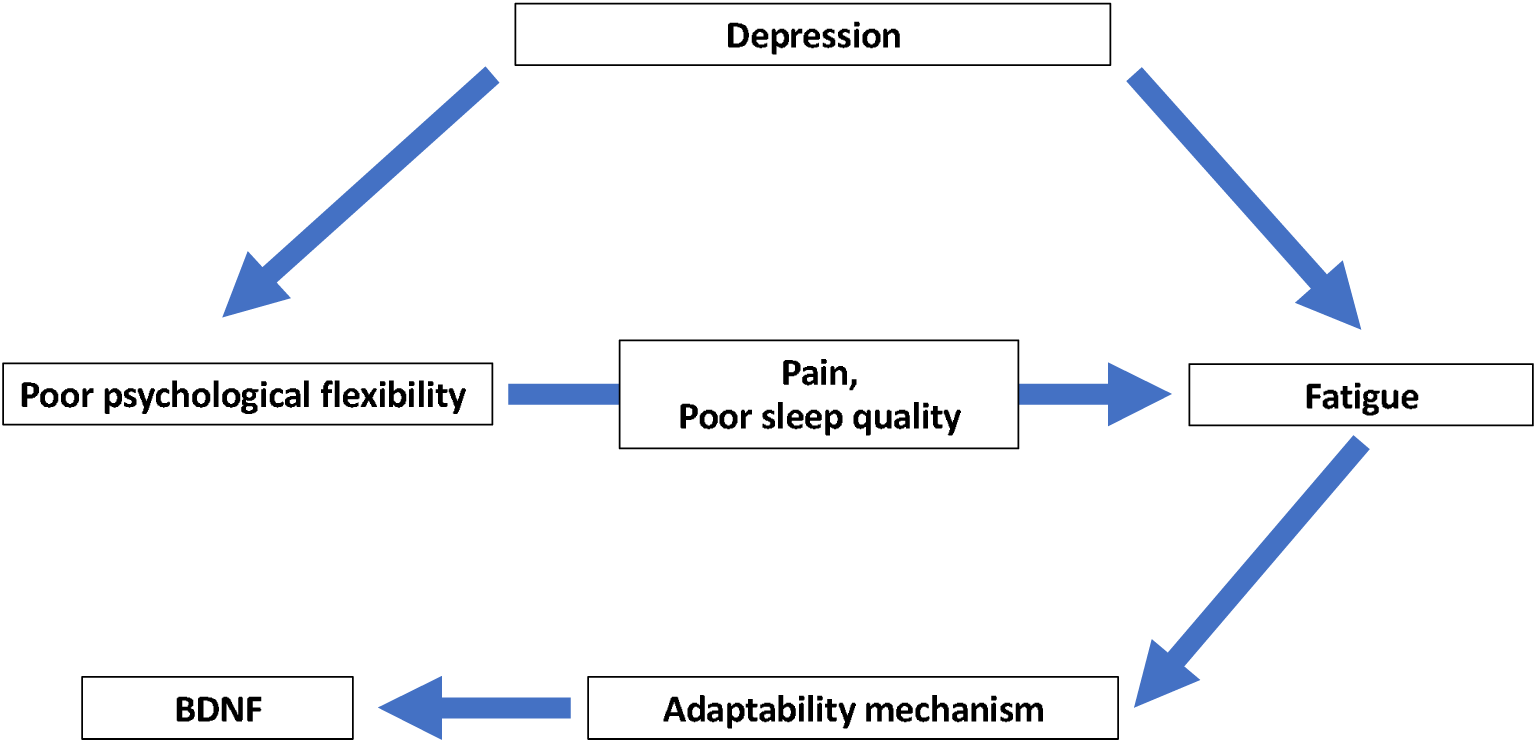
The association from our study data (Abbreviation: HRV = Heart Rate Variability, BDNF = Brain Derived Neurotrophic Factor)

Contrary to the previous study, our study HRV did not show a significant difference among people with fatigue and non-fatigue. This may be due to our volunteers all appearing healthy, fatigue group showing mild cases or are in the adaptive phase, in which physiological change may be too little to be detected by a relatively small sample size. The previous study showed that significantly lower HRV was associated with a higher level of fatigue symptoms(Fagundes et al., 2011). It could be noted that those studies were conducted with cancer patients therefore the difference in fatigue severity may be more obvious. Anyhow, our study shows a trend that the fatigue groups had lower HRV levels than non-fatigued groups in both the frequency domain and time domain that may be more significant in a larger sample size.

The morning serum cortisol in our study is also not significantly different between the fatigue and non-fatigue group. This issue may be explained for the same reason that a larger sample size may be needed to detect a small effect size in the mild fatigue group.(Golden et al., 2011)

There are some limitations to this pilot study, First, this study had a small sample size than detect a possible small effect size in early or mild fatigue people. Second, the cross-sectional study design inhibits the causative for the association between fatigue or psychological flexibility or high BDNF. Third, the short duration HRV rely on frequency domain like LF and HF may be not sensitive to minor physiological difference. The 24 hours HRV could use the time domain which may be more sensitive. Fourth, the negative psychological flexibility is a newly developed screening tool that needs repeated studying to prove their content validity. More studies should be conducted a larger samples, including the general population and other biomarkers such as DHEAs that should help detect and follow this condition.

## Conclusion

Our study supported the previous finding that low psychological flexibility and high BDNF were independent factors association with fatigue symptoms. This result provide insight that intervention that increase either psychological flexibility may prevent fatigue symptoms. The high BDNF may reflex the adaptive response of fatigue person and may be potential biomarkers for detecting early fatigue conditions. A larger sample size would be needed to approve this hypothesis.

## Data Availability

All data produced in the present study are available upon reasonable request to the authors

## Acknowledgements

The authors would like to thank Dr. James L. Wilson for his valuable advice and permission for some part of his questionnaire to be need in the psychological flexibility questions, and we would like to thank Associate Professor Tiam Srikhamjak for his advice on negative psychological flexibility questions for the reliability test.

## Author contributions

N.Y. and P.G. were responsible for the conception of the study, data collection, analyses, and interpretation of the data. N.Y. drafted the manuscript. N.Y. and P.G. critically revised the manuscript. All authors approved the final version of the manuscript and agree to be accountable in all aspects of the work.

## Notes

### Competing Interest Statement

The authors have declared no competing interest.

### Clinical Protocols

https://www.thaiclinicaltrials.org

### Funding Statement

None

### Author Declarations

Chiang Mai university's research ethics committee (FAM 2561 05328) gave ethical approveal for this work

### Summary of Updates

Revised the funding

